# Frequency and Potential Risk Factors Associated with the Development of Asymptomatic T2 Hyperintense Cervical Spine Lesions on MRI in Patients with Relapsing-Remitting Multiple Sclerosis

**DOI:** 10.1101/2024.09.24.24314299

**Authors:** Yasser Fadlallah, Yujie Wang, Muhammad Taimur Malik, Fan Tian, Peter A. Calabresi, Izlem Izbudak, Yishang Huang, Kathryn C. Fitzgerald, Ellen M. Mowry

## Abstract

**Background:** Multiple sclerosis (MS) is one of the most common non-traumatic neurologic disorders affecting young adults in the United States. Brain MRI is an important tool for monitoring disease activity and treatment efficacy in people with MS. Spinal cord (SC) imaging has been less consistently used for monitoring inflammatory disease activity, and the frequency of clinically silent breakthrough disease in the SC is still unknown. Given the particular vulnerability of the cervical spinal cord (c-SC) to inflammatory demyelination, it is important to evaluate the necessity of routine c-SC scanning or to ascertain appropriate strategies for its monitoring, considering the burden of each scan on a person with MS, including scanner discomfort and cost.

**Objective:** To determine how frequently follow-up magnetic resonance imaging (MRI) of the cervical spinal cord (c-SC) in patients with relapsing remitting multiple sclerosis (RRMS) reveals asymptomatic T2 hyperintense lesions, either in combination or in the absence of new MRI brain lesions, and to identify potential associated risk factors for developing such lesions.

**Methods:** Patients aged 18-65 years who were diagnosed with RRMS and were seen in longitudinal follow-up at the Johns Hopkins MS Center from January 1, 2014, to December 1, 2019, with an MRI brain and C spine performed during that period, were included. The results of up to four c-SC scans were considered during this study period. Asymptomatic new lesions were identified as new hyperintense T2 lesions, with or without enhancement, observed on MRI during routine follow-up surveillance. Univariate and multivariable-adjusted logistic regression (sex at birth, age, race, and current disease modifying therapy [DMT] category) were employed to identify factors associated with the development of an asymptomatic c-SC lesion for the first scan. Additionally, a mixed-effects logistic regression analysis was performed to assess factors associated with developing asymptomatic lesions across successive scans.

**Results:** A total of 869 individuals were included in the cohort, contributing a median of 3 (interquartile range: 2,4) scans per person. The proportion of incidental asymptomatic lesions identified ranged from 4.8 to 12.1% across the four scans analyzed in the study. Among those with new lesions in the c-SC, roughly half also showed concomitant new activity on brain MRI. The multivariate model was notable for Black/African Americans having higher odds of an asymptomatic new lesion (OR= 3.26, 95% CI 0.79, 5.93, p<0.0001), a result that persisted in mixed effects logistic regression models (OR = 1.73, 95% CI = 1.27, 2.35, p = 0.001). Higher-efficacy therapies were associated with higher odds of detection of a new lesion in the mixed-effects model, an association that was not present when considering just the first scan results as the outcome.

**Conclusion:** While the association of higher-efficacy therapy is presumed to be related to confounding by indication, Black/African American individuals with MS appear to be at higher risk of developing an asymptomatic c-SC lesion on MRI surveillance, which could suggest higher value of ordering asymptomatic screening imaging of the cord in this subpopulation. Ultimately, however, a very small percentage of the overall population of those with MS has a new cord lesion in the absence of symptoms, and half of those have a new lesion on screening brain MRI. These findings should motivate the creation and validation of predictive models to inform the utility cord imaging at a given timepoint for a given individual with MS, which could enhance healthcare quality and reduce costs.

## Introduction

Multiple sclerosis (MS) is a chronic autoimmune disorder of the central nervous system (CNS) that is characterized by inflammation, demyelination, and subsequent neuronal injury and degeneration [1]. MS is among the most prevalent causes of non-traumatic disability in young adults in developed countries [2]. Approximately 2.8 million people globally are thought to have MS [3]. The symptoms and unpredictable course of MS can significantly impact patients’ quality of life, affecting not only physical abilities but also emotional, psychological well-being, and various sociodemographic factors [4].

Magnetic resonance imaging (MRI) is regarded as the most important and reliable tool for diagnosing and monitoring MS patients due to its high sensitivity in detecting inflammatory changes in the brain and spinal cord [5]. Consequently, brain and spinal cord MRI have been integrated into the diagnostic criteria for MS, alongside clinical evaluation and cerebrospinal fluid (CSF) analysis, as outlined in the McDonald criteria, last updated in 2017 [6].

Periodic brain MRI is also extensively utilized for assessing response to treatment, even in the absence of new symptoms. According to the current Magnetic Resonance Imaging in MS (MAGNIMS)-Consortium of MS Centers (CMSC)-North American Imaging in MS Cooperative (NAIMS) guidelines, regular spinal cord imaging is not recommended in clinically stable patients with MS. This recommendation is partly due to the technical challenges and susceptibility to artifacts in spinal cord MRI, which can compromise image quality and offer limited additional benefit compared to brain MR. Imaging the thoracic spinal cord is particularly challenging due to more inconsistencies in imaging quality and susceptibility to artifacts [12]. Prior studies have suggested that spinal cord MRI offers limited additional benefit compared to brain MRI [8-9].

The presence of new asymptomatic brain MRI lesions detected during routine surveillance is associated with a greater likelihood of relapses and disease progression in MS patients [10]. Recent studies indicate that asymptomatic spinal cord lesions can occur independent of asymptomatic brain lesions, and that adding spinal cord MRI to routine surveillance might provide possible prognostic insight [11-12]. However, the impact of missing asymptomatic cord lesions on the disease course is yet to be clarified.

In this study, we aimed to assess the frequency of new, asymptomatic cervical spinal cord lesions detected via surveillance MRI in a group of clinically stable RRMS patients, with or without new asymptomatic brain lesions on MRI. Additionally, we sought to evaluate any potential clinical factors associated with the development of these lesions.

## Methods

### Study Population and Data Collection

We evaluated patients aged 18-65 years who were seen at the Johns Hopkins MS Center from January 1, 2014 to December 1, 2019. Patients were identified as presenting for routine follow-up surveillance, without new symptoms concerning for relapse, or due to symptoms, based on chart review by a qualified neurologist (Y.W. and M.T.M.). Symptomatic cases were defined clinically by the presence of newly developing neurological symptoms or the worsening of preexisting neurological dysfunction lasting for at least 24 hours, in the absence of fever or infections, and occurring at least 30 days after the preceding episode [13]. In contrast, asymptomatic patients presenting for routine surveillance were considered clinically stable.

### Radiological Assessment

During the study period, we retrospectively categorized scans as either conducted within the Johns Hopkins Health System (JHHS) or performed externally, with all scans re-reviewed by a JHHS radiologist and the reports generated at JHHS. We focused on the first four scans, labeled Scans 1-4, which correspond to the sequential cervical spine MRIs reviewed for each patient in the included cohort. Scan 1 refers to the initial scan recorded during the study period, which could have been for follow-up or symptomatic purposes. Subsequent scans (Scans 2, 3, and 4) are the next recorded MRIs for each patient, with the timing influenced by clinical indications (for surveillance vs symptoms) noted in the patient’s chart, rather than fixed intervals. We focused on asymptomatic patients, and we reviewed the corresponding radiology reports to identify the presence of new hyperintense T2 lesions and, if present, whether these lesions enhanced on post-contrast imaging. For the subgroup of individuals who had concomitant brain MRI, defined as having had such a scan within 6 months of the surveillance c-SC MRI, we also recorded whether the brain MRI demonstrated new lesions, since some clinicians might recommend a change in MS therapy if new activity was recorded on brain MRI, regardless of whether the cord also had new silent lesions, making the concomitant cord MRI less useful.

### Statistical Analysis

The primary outcome was defined as the presence or absence of an asymptomatic c-SC lesion detected on MRI. For patients presenting for initial cervical spine MRI (Scan 1), we used univariate logistic regression to compare baseline characteristics between patients with and without asymptomatic c-SC lesions. A multivariable logistic regression model, which included sex at birth, age, race, and current disease modifying therapy (DMT) category, was also employed to identify factors associated with the development of asymptomatic spinal lesions. These covariates were selected based on their clinical relevance and the rationale that they could influence MS lesion development. For the current DMT category variable, patients were initially grouped into moderate-efficacy DMT (glatiramer acetate, interferons, teriflunomide, dimethyl fumarate, fingolimod) and higher efficacy DMT (ocrelizumab, rituximab, natalizumab, alemtuzumab); those using no or unknown DMT were removed from multivariate analyses due to the low number in each group.

A mixed effects logistic regression model was used to longitudinally evaluate the factors associated with detecting asymptomatic c-SC lesions across successive scans during the study period. Additionally, we matched the first four cervical spine MRIs with contemporaneous brain MRIs, defined as those performed within 6 months of the spinal cord imaging. We then calculated the number needed to scan (NNS) to detect one asymptomatic c-SC lesion, with or without a concomitant brain lesion, across successive scans. Stata^®^ Statistical Software: Release 18 (College Station, TX: StataCorp LLC) and R software were used for analysis.

## Results

A total of 1,144 patients were identified as possibly eligible; after excluding progressive MS, clinically isolated syndrome, radiologically isolated syndrome, and other demyelinating diseases, 869 patients with RRMS who had brain and c-SC MRI were included in the analysis. Figure 1 illustrates the number of patients scanned at each point (Scan 1-4), the number who were asymptomatic at the time of the scan and thus included in the analyses, and the corresponding lesions detected on those follow-up scans. Among the cohort, 25.09% had one follow-up scan, 25.20% had two, 20.02% had three, and 16.46% had four follow-up scans. The cohort contributed a median of 3 (interquartile range (IQR): 2,4) scans per person.

**Figure 1.**
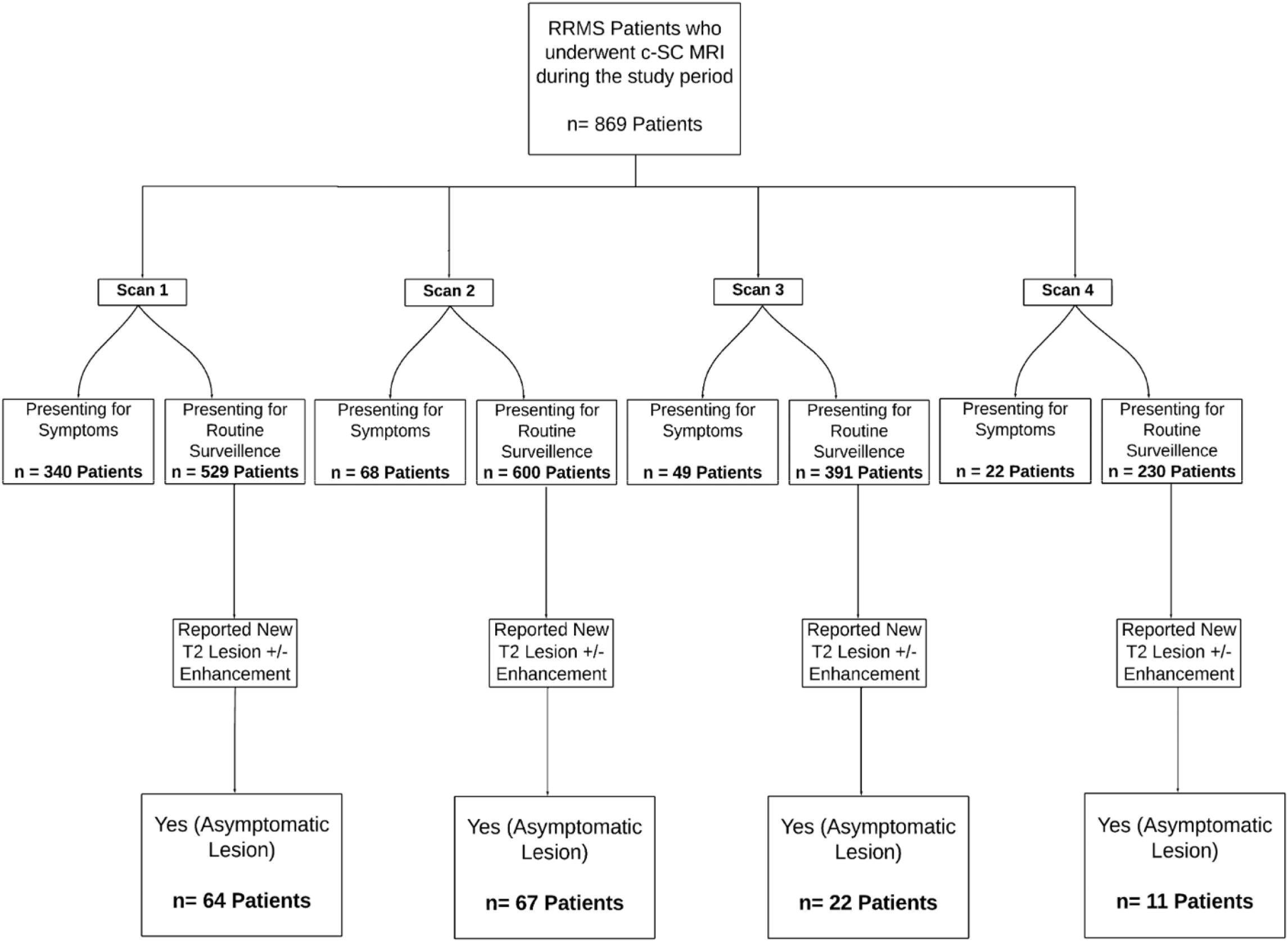
Flowchart of the Identification Process for Asymptomatic Lesions in RRMS Patients Undergoing Sequential Cervical Spinal Cord MRI (n = 869)

For the 869 patients included in the study, the baseline characteristics are summarized in Table 1. The mean age was 38.2 (standard deviation [SD] 7.3 years) with a female-to-male ratio of 3.4:1. The cohort was predominantly white (590, 67.9%), with 192 (22.1%) identifying as Black/African American. The mean symptom duration was 5.9 years (SD 5.5), and the median EDSS score was 1.5 (IQR 1.0, 2.5).

**Table 1.** Baseline Characteristics of the RRMS Cohort Examined During the Study Period.

|  |  |
| --- | --- |
| **Variable** |  |
| **N (number)** | 869 |
| **Age, years, mean (SD)** | 38.2 (7.3) |
| **Age, years** |  |
| Age <40 | 472 (54.32) |
| Age ≥ 40 | 397 (45.68) |
| **Sex** |  |
| Female, n (%) | 672 (77.3) |
| Male, n (%) | 197 (22.7) |
| **Race** |  |
| White, n (%) | 590 (67.9) |
| Black/African American, n (%) | 192 (22.1) |
| Other, n (%) | 87 (10.0) |
| **Symptom duration, years, mean (SD)** | 5.9 (5.5) |
| **Current disease modifying therapy (DMT)** |  |
| Not on DMT, n (%) | 29 (3.3) |
| Lower/Moderate-Efficacy Therapy, n (%) | 503 (57.9) |
| Higher Efficacy Therapy, n (%) | 311 (35.8) |
| Unknown, n (%) | 26 (3.0) |
| **Duration of use for current MS disease modifying therapy, months, mean (SD)** | 49.94 (36.1) |
| **Extended Disease Disability Scale (EDSS), median (Q1,Q3)** | 1.5 (1.0,2.5) |

In the univariate analysis comparing patients who did and did not develop asymptomatic cervical spinal cord lesions on their first MRI scan (Scan 1), Black/African American individuals had significantly higher odds of developing asymptomatic lesions (OR 2.88, 95% CI 1.64, 5.07, p < 0.0001). This finding persisted in the multivariate analysis after adjusting for age, sex, and disease-modifying therapy, with Black/African American patients maintaining increased odds of developing asymptomatic cervical cord lesions on Scan 1 (Table 2; OR 3.26, 95% CI 1.79–5.93, p < 0.0001).

**Table 2.** Multivariate Logistic Regression Analysis of Factors Associated with the Presence of Asymptomatic Cervical Spinal Cord Lesions on First Scan.

| Variable | Odds Ratio (95% CI) | p-value |
| --- | --- | --- |
| **Age** | 1.03 (0.99, 1.08) | 0.167 |
| **Sex** |  |  |
| Female | Reference |  |
| Male | 0.99 (0.52, 1.91) | 0.988 |
| **Race** |  |  |
| White/Caucasian-American, n (%) | Reference |  |
| Black/African American | 3.26 (0.79, 5.93) | <0.0001 |
| Other | 1.23 (0.41, 3.72) | 0.711 |
| **Current disease modifying therapy\*** |  |  |
| Lower/Moderate-Efficacy Therapy | Reference |  |
| Higher Efficacy Therapy | 0.80 (0.45, 1.44) | 0.464 |

\* No DMT' and 'Unknown' categories were excluded from this analysis due to low patient numbers, to ensure statistical robustness and reliability of the results.

Table 3 provides a longitudinal overview of asymptomatic cervical spinal cord (c-SC) lesions detected on MRI in RRMS patients across four time points. It shows the number of patients who underwent follow-up imaging, the percentage with asymptomatic c-SC T2 lesions, the presence of gadolinium enhancement, and the number needed to scan (NNS) to detect one asymptomatic T2 c-SC lesion. The number of patients imaged decreased over time, as did the percentage of new asymptomatic lesions, while the NNS increased from 8 at Scan 1 to 21 at Scan 4. Table 4 shows the relative yield of brain and c-SC asymptomatic lesions for each of the four matched c-SC and brain MRI scans. Asymptomatic new lesions were more common in the brain than in the c-SC. Among those who did have new lesions in the c-SC, these were accompanied roughly half of the time by concomitant new activity on brain MRI, as illustrated in Figure 2. The NNS to detect isolated new activity in the c-SC (in the absence of new brain MRI lesions) was higher later in the follow-up period (n=55 for Scan 4, versus n=18 for Scan 1).

**Table 3.**
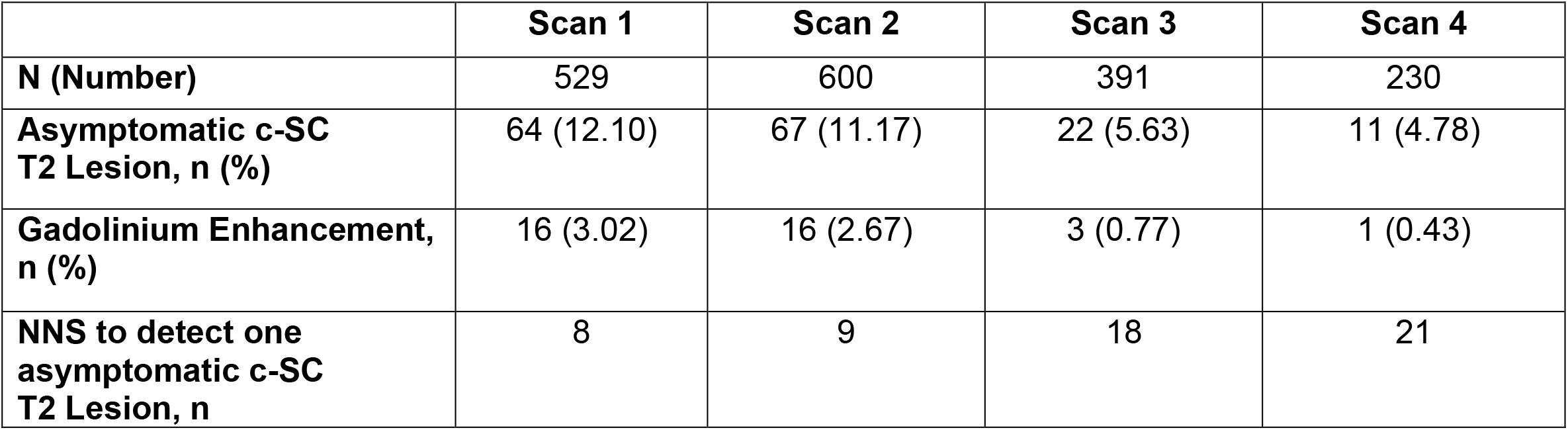
Longitudinal Overview of Asymptomatic Cervical Spinal Cord (c-SC) Lesions Detected on MRI in RRMS Patients and the Number Needed to Scan (NNS) to Detect One Asymptomatic T2 c-SC Lesion.

**Table 4.**
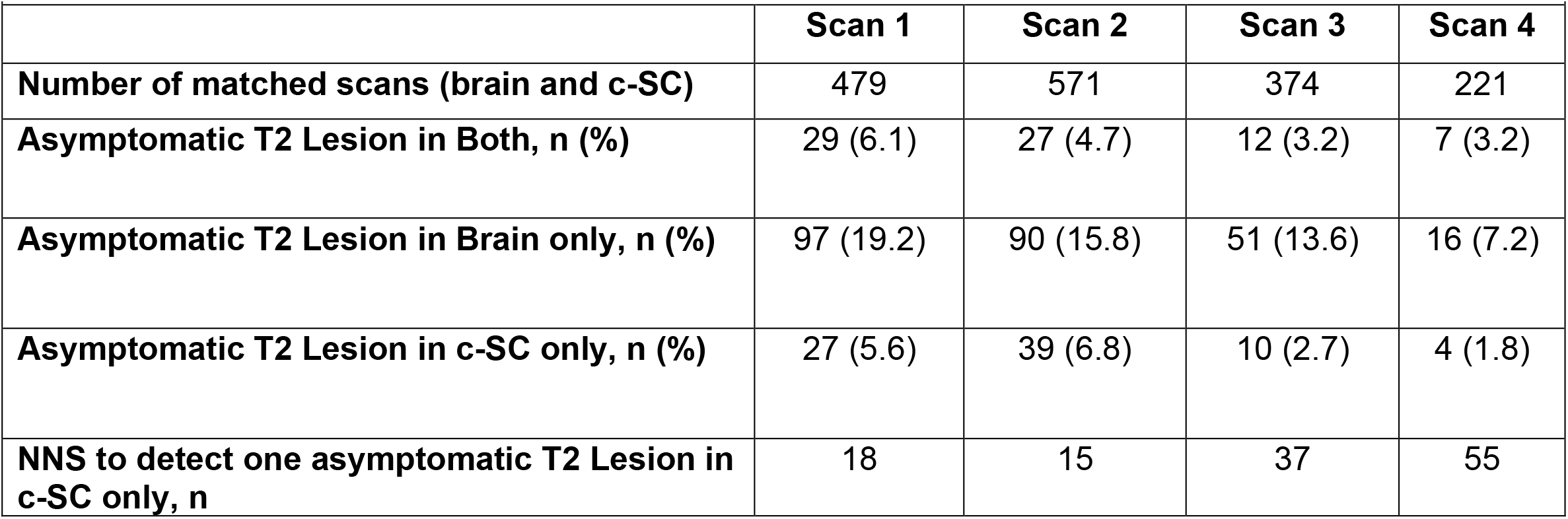
Relative Yield of Brain and Cervical Spinal Cord (c-SC) MRI in Detecting Asymptomatic New Lesion and the Number Needed to Scan (NNS) to Detect One Asymptomatic T2 c-SC Lesion Only (No Concomitant Asymptomatic Brain Lesion) in patients with RRMS.

**Figure 2.**
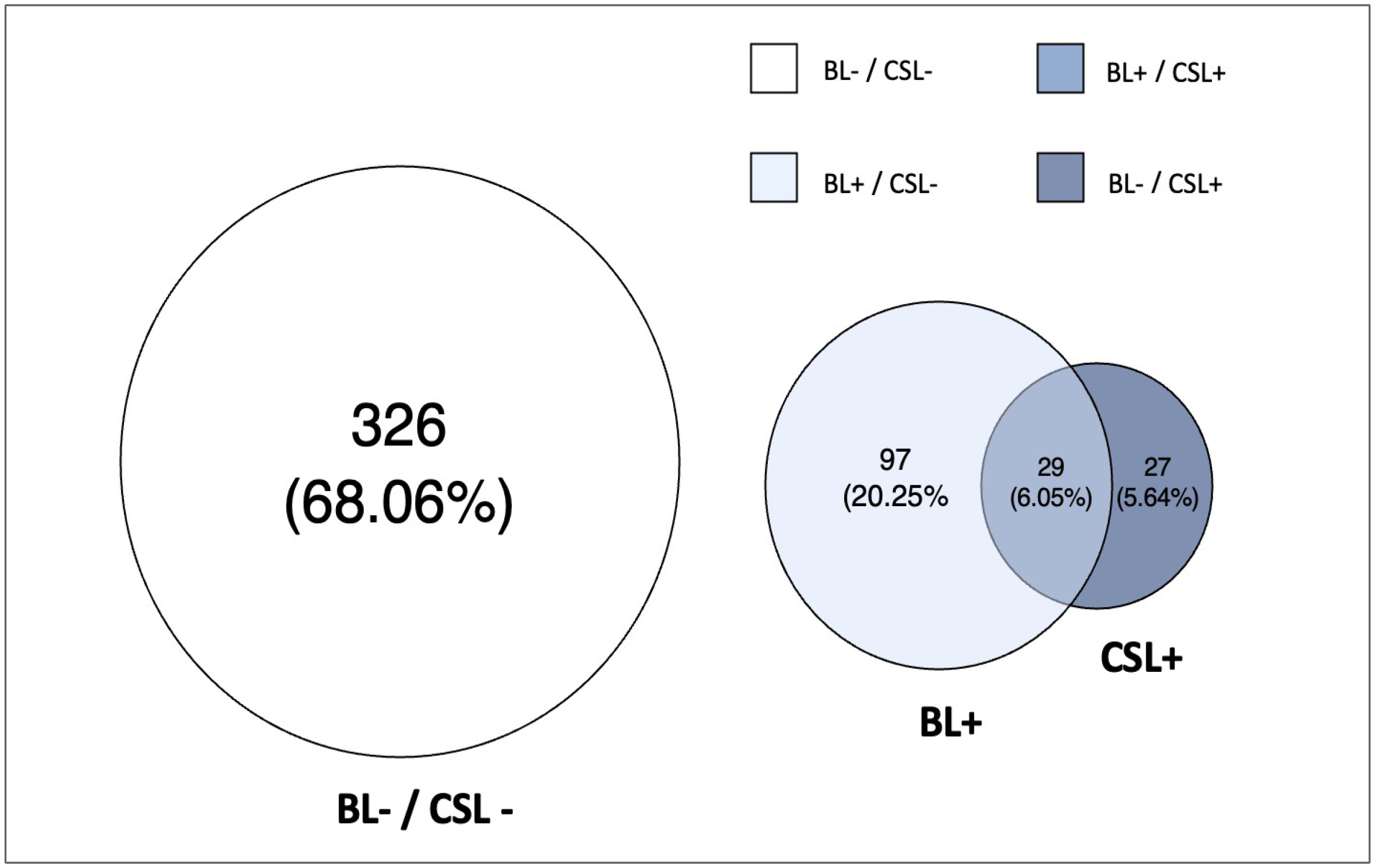
Frequency of patients with and without new lesions on MRI on First Matched brain and cervical spinal cord MRI Scan. BL−: No new asymptomatic lesion on brain MRI. BL+: At least one new asymptomatic lesion on brain MRI. CSL−: No new asymptomatic lesion on cervical spinal cord MRI. CSL+: At least one new asymptomatic lesion on c-SC MRI.

Finally, a mixed effects logistic regression was employed to longitudinally evaluate factors associated with developing asymptomatic c-SC lesions on successive c-SC scans (Table 5). The analysis indicated that the odds of developing asymptomatic c-SC lesions significantly decreased across the successive scans compared to Scan 1 (Scan 2 OR = 0.31, 95% CI = 0.23, 0.42; Scan 3 OR = 0.19, 95% CI = 0.13, 0.28; Scan 4 OR = 0.19, 95% CI = 0.12, 0.34; all p < 0.0001). Black/African American individuals continued to have greater odds of developing asymptomatic c-SC lesions compared to whites (OR 1.73, 95% CI 1.27, 2.35, p = 0.001) after adjustment for relevant covariates. Those using higher efficacy DMT had greater odds of developing asymptomatic lesions compared to patients receiving moderate-efficacy DMT (OR 1.56, 95% CI 1.19, 2.03, p = 0.001). This result remained significant even after excluding the first MRI scans conducted following a switch from lower to higher efficacy DMT (excluding 118 patients who had undergone DMT escalation to minimize confounding by indication (OR associated with use of higher-efficacy therapy= 1.44, 95% CI 1.07, 1.96, p = 0.017). Both Black/African American race and higher-efficacy therapy were associated with greater odds of an asymptomatic new lesion in sensitivity analyses in which only scans completed at Johns Hopkins were included.

**Table 5.** Longitudinal Analysis of Factors Influencing the Detection of Asymptomatic Cervical Spinal Lesions in RRMS patients Using Mixed Effects Logistic Regression.

| Variable | Odds Ratio (95% CI) | p-value |
| --- | --- | --- |
| **Scan** |  |  |
| Scan 1 | Reference |  |
| Scan 2 | 0.31(0.23, 0.42) | <0.0001 |
| Scan 3 | 0.19 (0.13, 0.28) | <0.0001 |
| Scan 4 | 0.19 (0.12, 0.34) | <0.0001 |
| **Age (per year)** | 1.00 (0.98, 1.02) | 0.783 |
| **Sex** |  |  |
| Female | Reference |  |
| Male | 0.92 (0.67, 1.27) | 0.622 |
| **Race** |  |  |
| White | Reference |  |
| Black/African American | 1.73 (1.27, 2.35) | 0.001 |
| Other | 1.35 (0.87, 2.09) | 0.186 |
| **Current disease modifying therapy\*** |  |  |
| Moderate Efficacy Therapy | Reference |  |
| Higher-Efficacy Therapy | 1.56 (1.19, 2.03) | 0.001 |

\* No DMT' and 'Unknown' categories were excluded from this analysis due to low patient numbers, to ensure statistical robustness and reliability of the results.

## Discussion

Although the value of spinal cord imaging in diagnosing MS is well-established, its utility in monitoring disease activity and assessing treatment response in clinical practice remains uncertain. While the 2021 MAGNIMS-CMSC-NAIMS consensus recommendations highlight that spinal cord MRI can be valuable for diagnosis, prognosis, and monitoring disease activity, the guidelines stop short of definitively recommending routine spinal cord MRI for monitoring clinically stable MS patients [7]. Herein, we found that the occurrence of asymptomatic c-SC lesions was rare, especially in the absence of new brain MS lesions, but was greater in Black/African American individuals with MS.

The incidence of asymptomatic c-SC lesions in clinically stable RRMS patients varies across studies. A recent study reported a 5.7% incidence over a median follow-up period of 14 months, [14] which is close to the 4.78% incidence observed in our study by Scan 4. In contrast, an older investigation, which included all spinal segments (not only the cervical segment), reported higher rates of asymptomatic spinal cord lesions, with 60% of MRI changes occurring asymptomatically [15]. Another group reported a 15% incidence of asymptomatic spinal cord lesions over a median period of 14 months in a cohort that included both RRMS and progressive MS patients [10]. The differences in findings between these studies and ours may be attributed to variations in study populations, technical factors (such as scanner type and magnetic field strength of the MRI scanner), imaging protocols used, methods of lesion confirmation, timing of scan relative to initiation or escalation of DMT, and DMT adherence.

While new asymptomatic c-SC lesions are less frequent than brain lesions, they can still occur independently, with up to a 15% incidence reported in stable MS patients [10-11]. In our cohort, 5.6% of patients had isolated asymptomatic c-SC lesions–higher than the 1.9% reported by Lim et al. (2024) but lower than the 10.12% found by Ostini et al. (2021) and the 8% observed by Lorefice et al. (2024) [10, 14, 16]. Additionally, approximately half of our patients with new asymptomatic c-SC lesions also had concomitant asymptomatic brain lesions, which suggests limited added diagnostic value of spinal cord imaging. Similarly, Tummala et al. (2017) demonstrated that combining spinal cord MRI with brain MRI for detecting “No Evidence of Disease Activity” (NEDA) in MS patients yielded minimal additional diagnostic value [17]. These findings support current consensus recommendations that routine brain MRI alone may be sufficient for detecting silent disease activity in clinically stable MS patients [10-11, 16].

In this study, we also found that Black/African American individuals were more likely to develop asymptomatic c-SC lesions. Previous studies have shown that Black/African American MS patients may have a more aggressive disease course and higher lesion burden compared to Caucasian patients, even after adjusting for socioeconomic status indicators, suggesting that factors beyond SES, such as other types of systemic racism, may contribute to these disparities [18,19, 20].

In addition, we found that patients receiving higher efficacy DMTs were more likely to develop asymptomatic c-SC lesions. While this finding at first seems counterintuitive, it is very likely due to confounding by indication, as patients on higher efficacy DMTs might have been prescribed these treatments due to a recent history of more active disease. We attempted to mitigate this concern by performing sensitivity analyses removing those who had escalated therapy, but it is likely that residual confounding is at play, considering it is known that higher-efficacy therapies are associated with reduced risk of new inflammatory lesions in randomized trials [21, 22, 23].

Our study has several limitations that should be acknowledged. First, the observational design limits the ability to infer causality, and the findings may be influenced by unmeasured confounding by indication of DMTs. We exclusively relied on existing radiological reports to determine the occurrence of lesions. This approach may present a challenge, as discrepancies can arise between the treating clinician’s interpretation and the radiologist’s report. However, this method reflects the reality of care for many patients, where treating clinicians often rely on radiologist interpretations due to lack of access or time to interpret cord MRI images directly. Further, clinicians’ interpretations can be influenced by their knowledge of the patient’s history and exam, while radiologists typically only know the diagnosis of MS. We intentionally focused on scans that had been reviewed by our institution’s radiologists to inconsistencies in lesion detection and reporting, but it is possible that this led to selection bias in the cohort, and our tertiary care center MS population is not likely fully representative of the broader MS population. The reliance on routine clinical MRI scans may also introduce variability in imaging quality and interpretation.

In conclusion, our study highlights that while the yield of surveillance imaging of the c-SC is relatively low in clinically stable RRMS patients, particularly when a concomitant brain MRI scan is free of new lesions, Black/African American individuals may be at a higher risk of developing asymptomatic c-SC lesions. To reduce out of pocket expenses and scan time for people with MS, clinicians may consider sequential scanning, ordering first a surveillance brain MRI and, only if that does not show new lesions, obtaining imaging of the c-SC. Our findings also highlight an opportunity to develop predictive models to determine the likelihood of detecting a new lesion at a given time point for a person with RRMS. Such models could be incorporated into a clinical decision support (CDS) tool to enable decision-making by treating clinicians and patients surrounding whether to acquire c-SC imaging to monitor MS activity over time.

## Data Availability

All data produced in the present study are available upon reasonable request to the authors.

